# Clinical evaluation of the Roche distributed SD Biosensor SARS-CoV-2 & Flu A/B Rapid Antigen Test amongst mild symptomatic people during the 2022/2023 winter season

**DOI:** 10.1101/2024.02.05.24302338

**Authors:** Zsὁfia Iglὁi, Jans Velzing, Marion Koopmans, Richard Molenkamp

**Affiliations:** Erasmus MC, Rotterdam, The Netherlands

## Abstract

Both influenza and SARS-CoV-2 are seasonal respiratory illnesses with similar symptoms, however distinguishing one from the other can have benefits for the patient and have different implications in various settings.

In this study we have evaluated the clinical performance of the Roche distributed SD Biosensor SARS-CoV-2 & Flu A/B Rapid Antigen Test during the 2022/2023 winter season, in a non-hospitalized, mild symptomatic population, comparing results with reverse transcription quantitative polymerase chain reaction (RT-qPCR). Participants also filled in a short questionnaire about their symptom onset, symptoms, vaccination status for both influenza and SARS-CoV-2.

We could include 290 people with complete records with female majority (72%, 209/290). Age ranged from 18 years old (minimum age for inclusion) to 71 years (mean age was 40.4 years). From the 290 inclusions 93 tested positive with SARS-CoV-2 PCR, 12 by influenza A and 6 by influenza B PCR. For SARS-CoV-2 overall sensitivity was 72.0% (confidence interval, CI 61.8-80.9%) and specificity 99.5% (CI 97.2-99.9%). SARS-CoV-2 RDT performed best up to and including PCR ct value of 25 (sensitivity 96% CI 85.8-99.5%), but could also detect samples less or equal to PCR ct 33, however with lower sensitivity (sensitivity 80.0% CI 69.6-88.1%). For influenza limited amount of samples were available; the RDT detected influenza A with 58.3% sensitivity (CI 27.7-84.8) and 100% specificity (CI 98.6-100.0%). In case of influenza B the inclusions were too low to calculate sensitivity reliably (2/6, 33.3% CI 4.3-77.7%); specificity was 98.2% (5/274, CI 95.8-99.4%). No cross reaction between SARS-CoV-2 and Flu A/B was experienced.

As was shown before, SARS-CoV-2 could be determined with high sensitivity in recent onset and lower than ct 25 samples. In spite of performing the study throughout the influenza season, we had sub optimal inclusions for determining RDT clinical performance; further studies are needed.

## 1. Introduction

Antigen rapid tests are very useful tools to identify the causative agent rapidly on the spot. Influenza is an important human pathogen which is with humanity since centuries and caused several pandemics [1]. In 2019 SARS-CoV-2 emerged and caused the largest pandemic of modern times. Life is now back to normal, but looks like SARS-CoV-2 is here to stay as a seasonal respiratory illness.

For the year of (2022/2023) a strong and early influenza season was anticipated due to the low circulation in the previous 2 years [2]. Given that both viruses are respiratory viruses and display similar symptoms, the chance that people with respiratory symptoms are in fact infected with influenza rather than SARS-CoV-2 is anticipated to be high during the influenza season.

Infection by one or the other virus might have different implications for the patients especially in healthcare settings, therefore distinguishing them as soon as possible would be beneficial. Numerous diagnostic tests exist for influenza including point of care test utilizing reverse transcription quantitative polymerase chain reaction (POCT RT-qPCR), however currently there is no reliable influenza antigen RDT on the market [3]. Antigen rapid tests (RDTs) became part of standard diagnostic test repertoire for SARS-CoV-2 in numerous countries and various settings. Using RDT can be beneficial especially in settings like nursing homes, schools, workplaces etc. In hospitals and for vulnerable population molecular POCT are preferred. In preparation for future seasonal co-circulation of these two respiratory viruses, availability of reliable rapid diagnostic tests which can detect both of these viruses simultaneously is necessary. In this study we have evaluated the clinical performance of the Roche distributed SD Biosensor SARS-CoV-2 & Flu A/B Rapid Antigen Test during the 2022/2023 winter season, in a non-hospitalized, mild symptomatic population, to test future feasibility for use. RDT results were compared to RT-qPCR results as gold standard method for both viruses. Symptoms and date of symptom onset was collected. Vaccination status for both SARS-CoV-2 and influenza, date and type of vaccine was asked in a short questionnaire.

## 2. Methods

### 2.1 Testing population and patient recruitment process

Employees of the Erasmus Medical center, with or without symptoms, were eligible for free of charge PCR testing up until 1^st^ April 2023 (due to policy change, testing for healthcare workers was no longer required). Appointments for testing were arranged via a call center serving specifically the test center. Participants were recruited during this phone call, which also enabled forward planning of the amount of tests/day. Participants signed the informed consent and filled in the short questionnaire during their appointment. We started inclusion on 15^th^ December 2022 (start of the influenza season) and continued till 30^st^ March 2023 with the intention of catching the peak of both influenza A and B. We only recruited symptomatic individuals.

### 2.2 Specimen collection and testing procedures

Standard method for SARS-CoV-2 and influenza testing is by RT-qPCR which was carried out as usual, in parallel with the RDT. One combined swab (oro- and nasopharyngeal swab, OP + NP swab) was taken for RT-qPCR, placed directly in universal transport media (HiViral™) and testing was performed using the Aptima™ SARS-CoV-2/Flu Assay (Panther® System – Hologic). Please note that for the influenza PCR no ct values are available only Transcription Mediated Amplification (TMA) values. For the RDT evaluation, a second NP swab was taken from the same or the other nostril using the swab included in the kits to directly compare RT-qPCR result with the RDT. Test was performed within the manufacturer recommended time (<30mins) following instructions.

### 2.3 Data analysis

Results from the RDT and questionnaire were collected in Microsoft Access. Results from the PCR were merged together with this. Sensitivity and specificity of the RDT compared to the RT-qPCR results were calculated for the whole dataset and also for specific subsets. Clopper-Pearson analysis was be used to determine confidence intervals of proportions. Two sample t-test was used to define significance of difference between means. R version 4.0.2 was used to merge, clean and analyze the data.

### 2.4 Ethical clearance

Ethics committee of Erasmus MC, Rotterdam, The Netherlands waived ethical approval for this work (protocol number MEC-2021-0943).

## 3. Results

### 3.1 Characteristics of included population

In total we had 290 complete patient data set available at the end of the study with female majority (72%, 209/290). Age ranged from 18 years old (minimum age for inclusion) to 71 years (mean age was 40.4 years; sex specific mean age: males 41.9 vs female 39.7 years); dominant age group was the 28-37 years (29%, 83/290), followed by the 38-47 years old (21%, 62/290) and the 18-27 years old (19%, 56/290) (Table 1). Since the presence of symptoms was inclusion criteria, all participants claimed to be symptomatic and majority (81%, 236/290) had recent onset i.e. less than 7 days. Most common symptoms amongst SARS-CoV-2 PCR positive participants were runny nose (76/93), cough (64/93), throat ache (63/93), tiredness (41/93), headache (39/93), myalgia (27/93), productive cough (23/93), breathlessness (19/93), cold chills (19/93), fever (17/93). Nausea, diarrhea, eye pain, painful breath, swollen lymph nodes, vomiting or nosebleed was reported in very few cases; rash wasn’t reported. Most common symptoms amongst influenza A PCR positive participants were cough (10/12), runny nose (8/12), headache (8/12), throat ache (6/12), cold chills (6/12), fever (5/12), myalgia (5/12), tiredness (4/12), breathlessness (3/12), eye pain (3/12). Productive cough, diarrhea, painful breath was reported in very few cases; nausea, rash, vomiting, swollen lymph nodes or nosebleed wasn’t reported.

**Table 1.** Age and sex characteristics of the included participants.

| Age categories | Females | Proportion females/age category | Males | Proportion males/age category | Proportion of total (n=290) | Total |
| --- | --- | --- | --- | --- | --- | --- |
|  | No. | % | No. | % | % | No. |
| **18-27** | 42 | 75% | 14 | 25% | 19% | 56 |
| **28-37** | 62 | 75% | 21 | 25% | 29% | 83 |
| **38-47** | 45 | 73% | 17 | 27% | 21% | 62 |
| **48-57** | 37 | 80% | 9 | 20% | 16% | 46 |
| **58-67** | 23 | 59% | 16 | 41% | 13% | 39 |
| **68-71** | 2 | 50% | 2 | 50% | 2% | 4 |
| **Total** | 211 | 73% | 79 | 27% | / | 290 |

### 3.2 Performance of the SARS-CoV-2 *Ag RDT*

In total 290 samples were tested by both PCR and RDT and 32% (93/290) was PCR positive. Majority of samples were in the PCR ct 18-33 range (86%, 80/93) and had recent onset i.e. <7 days (89%, 81/91 of known onset). Overall sensitivity was 72.0% (67/93, confidence interval, CI 61.8-80.9%) and specificity 99.5% (1/197, CI 97.2-99.9%). The RDT performed best under and including PCR ct value of 25 (44/46, sensitivity 96% CI 85.8-99.5%) but could detect samples less or equal to PCR ct 33 with lower sensitivity (64/80, sensitivity 80.0% CI 76.6-88.1%). Samples with lower ct values had more recent onset than the ones in higher ct categories but nevertheless all samples with < PCR ct 41 were <7 days since symptom start (Table 2).

**Table 2.** Overview of the SARS-CoV-2 specific results by PCR ct values, median days since onset with IQR and RDT positivity.

| PCR ct categories | Median days since onset | Interquartile range (IQR) | Total PCR positives | Total PCR negatives | Total RDT positives | Total RDT negatives | Percentage RDT positives |
| --- | --- | --- | --- | --- | --- | --- | --- |
|  | Days | Days | No. | No. | No. | No. | % |
| **18-25** | 1 day | 1.0 | 46 | 0 | 44 | 2 | 96% |
| **26-33** | 1 day | 2.0 | 34 | 0 | 20 | 14 | 59% |
| **34-41** | 2 days | 6.5 | 12 | 0 | 2 | 10 | 17% |
| **42-44** | *na* | *na* | 1 | 0 | 0 | 1 | 0% |
| **45+** | 1 day | 2.0 | 0 | 197 | 1 | 196 | *na* |
| **Total** | / | / | 93 | 197 | 67 | 223 | / |

### 3.3 Performance of the Flu A/B *Ag RDT*

Only 12 influenza A PCR positive and 6 influenza B PCR positive samples were detected, which are both too low to calculate sensitivity reliably. Influenza A was detected with 58.3% sensitivity (7/12, CI 27.7-84.8%) and 100% specificity (0/269, CI 98.6-100%). In case of influenza B sensitivity is 33.3% (2/6, CI 4.3-77.7%) and specificity was 98.2% (5/274, CI 95.8-99.4) (Table 3).

**Table 3.** Overview of influenza A and B specific results by PCR and RDT results.

| Flu A PCR | Flu A RDT positive | Flu A RDT negative | Flu B PCR | Flu B RDT positive | Flu B RDT negative | Flu B RDT borderline |
| --- | --- | --- | --- | --- | --- | --- |
| Result | No. | No. | Result | No. | No. | No. |
| Negative | 0 | 269 | Negative | 2 | 269 | 3 |
| Positive | 7 | 5 | Positive | 2 | 4 | / |
| Total/category | 7 | 274 | Total/category | 4 | 273 | 3 |
| Total | 281 |  | Total | 280 |  |  |

### 3.4 Results in the context of vaccinations

Vast majority of the included people were vaccinated against SARS-CoV-2 (94%, 274/290), however time since vaccination varied and there is a decreasing proportion of participants who got vaccinated following the initial vaccination. The proportion positives are similar independently from the amount of vaccinations, however slightly higher in the group which received 4x vaccinations vs. less or more (Table 4). For influenza, half of the included participants were vaccinated in 2022 October/November against influenza (50%, 145/290). Similar proportions were testing positive (7/144 not vaccinated and 5/144 vaccinated).

**Table 4.** Results in light of vaccination.

|  | **Vaccinated 1x**<br>**(n) and**<br>**% of**<br>**total** | **Median**<br>**(min**<br>**max**<br>**and n)**<br>**days**<br>**since**<br>**vaccinat**<br>**ion** | **Vaccinated 2x**<br>**(n) and**<br>**% of**<br>**total** | **Median**<br>**(min**<br>**max**<br>**and n)**<br>**days**<br>**since**<br>**vaccinat**<br>**ion** | **Vaccinated 3x**<br>**(n) and**<br>**% of**<br>**total** | **Median**<br>**(min**<br>**max**<br>**and n)**<br>**days**<br>**since**<br>**vaccinat**<br>**ion** | **Vaccinated 4x**<br>**(n) and**<br>**% of**<br>**total** | **Median**<br>**(min**<br>**max**<br>**and n)**<br>**days**<br>**since**<br>**vaccinat**<br>**ion** | **Vaccinated 5x**<br>**(n) and**<br>**% of**<br>**total** | **Median**<br>**(min**<br>**max**<br>**and n)**<br>**days**<br>**since**<br>**vaccinat**<br>**ion** |
| --- | --- | --- | --- | --- | --- | --- | --- | --- | --- | --- |
| **SARS**<br>**-CoV**<br>**PCR**<br>**Positi**<br>**ve** | 83<br>(32%) | 640<br>(1075-232) | 82<br>(33%) | 593<br>(3680-33) | 65<br>(32%) | 403<br>(814-25) | 41<br>(39%) | 105<br>(725-4) | 5 (38%) | 108<br>(336-48) |
| **SARS**<br>**-CoV**<br>**PCR**<br>**Negat**<br>**ive** | 179<br>(68%) |  | 163<br>(67%) |  | 139<br>(68%) |  | 64<br>(61%) |  | 8 (62%) |  |
| **RDT**<br>**positi**<br>**ve** | 58<br>(22%) |  | 57<br>(23%) |  | 47<br>(23%) |  | 30<br>(29%) |  | 3 (23%) |  |
| **RDT**<br>**negati**<br>**ve** | 204<br>(78%) |  | 188<br>(77%) |  | 157<br>(23%) |  | 75<br>(71%) |  | 10<br>(77%) |  |
| **Total** | **262** | **244\*** | **245** | **228\*** | **204** | **191\*** | **105** | **95\*** | **13** | **12\*** |

## 4. Discussion

This study was carried to establish the clinical performance of this SARS-CoV-2 and influenza combination RDT. In spite of careful planning to cover the entire influenza season and thus include both influenza A and B, we could only achieve suboptimal inclusion, which could only partly establish the clinical performance of this test (SARS-CoV-2).

For SARS-CoV-2 the overall sensitivity of the RDT was 72.0%. This value is lower than what was originally detected by the earlier version of this test at the beginning of the pandemic [4], however it is in line with the trend what was noticed later in the pandemic [5]. Early days since symptom onset do not necessarily produce low PCR ct values anymore due to existing immunity and vaccination. This is also lowering the detected sensitivity of the RDTs which are most sensitive with high viral load, which was still the case in this study; sensitivity for <=PCR ct 25 was still 96%. Results are skewed towards females and younger age groups (<50 years of age), however the proportion positives are similar between sexes across most age groups (Table 1). Symptoms were not tested for statistical significance, some symptom were slightly more common amongst COVID-19 positive participants. However SARS-CoV-2 is still evolving thus the displayed symptoms change just like for influenza.

Vaccination in this study does not seem to influence the disease or the testing outcome as similar proportion were tested positive with RDT as with PCR and this was true across the whole group independently of the amount of vaccination received.

In summary, there is a clear benefit to have a combination test for commonly co-circulating seasonal viruses, however further studies are needed to establish the clinical performance of the influenza A and B part of this RDT.

## Data Availability

Aggregated data produced in the present study are available upon reasonable request to the authors

## 5. Acknowledgement

We would like to express our special thanks to the employees of the swab unit at Erasmus MC for their efforts to include participants in this study and the willingness to take on the extra work load what this represented. Furthermore we would like to thank all participants as without your contribution no research projects can be executed.

